# Public health information on COVID-19 for international travellers: Lessons learned from a rapid mixed-method evaluation in the UK containment phase

**DOI:** 10.1101/2020.09.22.20195628

**Authors:** Tinting Zhang, Charlotte Robin, Shenghan Cai, Clare Sawyer, Wendy Rice, Louise E. Smith, Richard Amlôt, G. James Rubin, Rosy Reynolds, Lucy Yardley, Matthew Hickman, Isabel Oliver, Helen Lambert

## Abstract

**Introduction:** In the containment phase of the response to the COVID-19 outbreak, Public Health England (PHE) delivered advice to travellers arriving at major UK ports. We aimed to rapidly evaluate the impact and effectiveness of these communication materials for passengers in the early stages of the pandemic.

**Methods:** In stage I (Patient and Public Involvement, PPI) we interviewed seven travellers who had returned from China in January and February 2020. We used these results to develop a questionnaire and topic guides for stage II, a cross-sectional survey and follow-up interviews with passengers arriving at London Heathrow Airport on scheduled flights from China and Singapore. The survey assessed passengers’ knowledge of symptoms, actions to take and attitudes towards PHE COVID-19 public health information; interviews explored their views of official public health information and self-isolation.

**Results:** In stage II, 121 passengers participated in the survey and 15 in follow-up interviews. 83% of surveyed passengers correctly identified all three COVID-19 associated symptoms listed in PHE information at that time. Most could identify the recommended actions and found the advice understandable and trustworthy. Interviews revealed that passengers shared concerns about the lack of wider official action, and that passengers’ knowledge had been acquired elsewhere as much from PHE. Respondents also noted their own agency in choosing to self-isolate, partially as a self-protective measure.

**Conclusion:** PHE COVID-19 public health information was perceived as clear and acceptable, but we found that passengers acquired knowledge from various sources and they saw the provision of information alone on arrival as an insufficient official response. Our study provides fresh insights into the importance of taking greater account of diverse information sources and of the need for public assurance in creating public health information materials to address global health threats.

**What is already known?:**

- In the containment phase, PHE issued public health advice at the major UK ports of entry to arriving travellers in response to the COVID-19 outbreak.

**What are the new findings?:**

- The majority of passengers correctly identified all three symptoms of COVID-19 highlighted in the PHE advice at the time and understood the importance of reporting symptoms and self-isolation.
- Knowledge about COVID-19 was also acquired elsewhere and was often more extensive than the information provided in official PHE guidance.
- PHE advice was perceived as clear and acceptable but insufficient on its own as an official response to the pandemic.

**What do the new findings imply?:**

- Our evaluation shows that while the PHE leaflets and posters met the intended aim of providing information and guidance, passengers used the provision of information and other visible public health measures to judge the adequacy of governmental response to the pandemic.
- Our study provides fresh insights into the need to take greater account of the diverse information sources from which international travellers may draw.
- Our study indicates that public health measures instituted at borders should be appraised not only with respect to their functional effectiveness in contributing to infection control, but also for their perceived effectiveness in furnishing public assurance of official action to contain the disease threat, which could be helpful in building public trust and thereby encouraging adherence to official guidelines.
- Our study demonstrates the complexity of health policy decision-making in public health emergencies of international importance and highlights the importance of establishing efficient mechanisms for rapid appraisal and feedback to public health and regulatory authorities of evidence that could contribute to containment and control of epidemic disease threats.

## INTRODUCTION

Large-scale frequent international population movements are occurring in our globalised societies, with international arrivals growing to 1.186 billion in 2015.[1] The increasing connectivity between countries has increased pressure on prevention and coantainment of global disease outbreaks. The current COVID-19 pandemic is a major threat which has profound and serious impacts on global public health. In December 2019, pneumonia cases of unknown aetiology were reported in Wuhan, China;[2] this illness was linked to a novel coronavirus on the 7^th^ of January 2020.[3] The first cases of COVID-19 in England were reported on 29^th^ January in two people who had recently arrived from China. Initial cases were mostly associated with international travel. As of the 13^th^ September 2020, there have been 368,504 positive cases in the UK.

Alongside pharmaceutical interventions, public health information is essential to controlling the COVID-19 pandemic.[4] Provision of public health advice at ports of entry, including the symptoms and actions to take if they develop, was most recently used in the UK in the context of the response to the Ebola outbreak in West Africa in 2014/15, which was appreciated and considered as reassuring for the public by travellers.[5] During the containment phase of the COVID-19 response, when the epicentre of the outbreak was in Asia, public health information was delivered to travellers arriving at UK ports up to the point when extensive travel restrictions were implemented. The Airport Public Health Monitoring Operations Centre established by Public Health England (PHE) was activated on 25^th^ January to monitor all direct flights from China to LHR, and operations were extended to include all direct flights to London Gatwick and Manchester on 29^th^ January. Measures directed at passengers travelling from affected countries into the UK included a broadcast message to passengers made on incoming aircraft, to encourage travellers to report their illness; posters containing COVID-19 related public health advice displayed at these three airports; and leaflets containing this advice provided to passengers by airlines on board the flight and/or made available on arrival. Up to 13^th^ March (the last data collection time point of this study), these leaflets and posters were updated three times, with key information summarised in Box 1. Contact tracing was undertaken when a case was reporting including flights and other transport. Since 8^th^ June, more restrictive rules have been instituted for incoming passengers: people who enter or return to the UK are required to provide their journey and contact details and self-isolate for 14 days if arriving from an affected country, with penalties of up to £1,000 for breaking this rule.[6] These regulations continue to be amended, with exemptions for travellers arriving from specified countries of origin.

The ongoing risk associated with travel highlights the importance of interventions that target arriving passengers to control transmission and protect the public. A previous study showed the importance of evaluating interventions delivered during international epidemics to inform the design of similar initiatives and to ensure their acceptability and adherence in the future.[5] The aim of this study was to evaluate the effectiveness and impact of PHE COVID-19 related communication materials for passengers arriving at UK airports during the containment phase of the response (24^th^ January to 12^th^ March). The study was conducted at the request of the Department of Health and Social Care via the National Institute of Health Research. Adjustments to the study protocol were made due to the fast-changing situation, as the number of flights carrying passengers into the UK dropped substantially in the monitoring period, from 16-18 flights per day from China (including Hong Kong) into LHR in the third week of January, to only 9 flights per week by the end of January, with air traffic reducing further in the subsequent months. Internal data from LHR indicated that in March a total of 123 flight arrived from China, Hong Kong and Singapore, one-fifth of the number in February.

**Box 1.** Key messages of leaflets and posters distributed at major ports of entry in England (in English and simplified Chinese)

| Version | Developed/<br>Updated Date | Case Definition | Advice |
| --- | --- | --- | --- |
| 1 | 23/Jan/2020 | A new infection has been detected in Wuhan, China<br>If you have been to Wuhan, China, in the last 14 days and develop ANY of these symptoms:<br>Cough, Sore throat, Difficulty breathing, Fever, Runny nose | Contact a healthcare professional<br>If you develop symptoms within 14 days of travel to Wuhan, please stay indoors and avoid contact with others where possible. Ring 111 or call your GP and tell them that you have travelled to Wuhan, for free advice and treatment |
| 2 | 31/Jan/2020 | If you have been to Wuhan, or Hubei Province, China in the last 14 days... | Stay indoors and avoid contact with others<br>Contact NHS 111 for advice<br>Please follow this advice even if you do not have symptoms of the virus<br>Do not go to work, school, or public areas<br>Avoid visitors in your home<br>Avoid using public transport or taxi |
|  |  | If you have been to anywhere on mainland China in the last 14 days and develop any of these symptoms<br>Cough, fever, shortness of breath | Stay indoors and call NHS 111 |
| 3 | 07/Feb/2020 | If you have a cough, or fever, or shortness of breath and have been to any of these places in the last 14 days:<br>China, Thailand, Japan, Republic of Korea, Hong Kong, Taiwan, Singapore, Malaysia, Macau | Stay indoors and call NHS 111 |
|  |  | If you have been to Wuhan, or Hubei Province, China in the last 14 days... | Stay indoors and contact NHS 111 even if you do not have symptoms |
| 4 | 25/Feb/2020 | If you have a cough, or fever, or shortness of breath and have been to any of these places in the last 14 days:<br>China, Macau, Thailand, Vietnam, Japan, Cambodia, Republic of Korea, Laos, Hong Kong, Myanmar, Taiwan, Northern Italy (north of Pisa), Singapore, Malaysia | Stay indoors and call NHS 111 |
|  |  | If you have returned from these specific areas:<br>Iran, Daegu or Cheongdo in South Korea, Specified towns in the Lombardy and Veneto regions in Northern Italy, Hubei province (Returned in the past 14 days) | Make your way to your residence. Stay indoors and contact NHS 111 even if you do not have symptoms |

Source from: https://campaignresources.phe.gov.uk/resources/search

## METHODS

We undertook a two-stage mixed-methods evaluation, starting with patient and public involvement (PPI) interviews with Chinese students and staff at two UK universities, followed by a survey and semi-structured interviews with air passengers returning to the UK from COVID-19 affected countries.

### Study population

Stage I: Staff and students from two universities in England who had returned to the UK from China in January and February 2020.

Stage II: Returning travellers aged 18 years and over from any nationality, arriving into LHR airport from affected countries after PHE leaflets and posters began to be distributed on 23^rd^ January.

## Sampling and methods

### Stage I – Patient and Public Involvement (PPI) interviews

Volunteers were recruited from King’s College London and University of Bristol though invitations posted on internal websites and e-lists and snowball sampling. One-to-one telephone interviews of approximately 30 minutes were conducted between 4^th^ and 14^th^ February 2020. Participants with recent experience of arriving from Asia were asked about what they had seen at the airport, and their comments on PHE information and advice about COVID-19 were elicited. To facilitate this, we provided participants with copies of the leaflets and posters being displayed at airports. The interviews were audio recorded; data from each interview was summarised and synthesised to build a final report. Based on the PPI results, a questionnaire and interview topic guides were developed for Stage II.

### Stage II – Airport cross-sectional survey and follow-up interviews

#### Cross-sectional survey

Passengers arriving into LHR airport, UK on three scheduled flights operated by three airlines on 4^th^ March from Singapore and on 12^th^ March and 13^th^ March from China, were recruited into the survey. During the study period, PHE information listed both these countries as places of origin that necessitated advice for travellers, with Hubei and Wuhan in China highlighted as special places of origin with separate advice. Instructions were provided to airline crews in advance. Paper questionnaires provided in English, Mandarin and Cantonese versions and the leaflets produced by PHE in English and simplified Chinese script were issued by crew to all passengers for completion prior to disembarkation. There were in total 11 questions included in the questionnaire to collect information on three main aspects, including: participants’ knowledge of COVID-19 symptoms (Q1) and help-seeking behaviours (Q2), whether participants received the public health advice (Q3) and their views on it (Q4), and their demographic information (Q5-Q11).

Respondents were also invited to record their name and contact details on the questionnaires if willing to take part in follow-up interviews. Researchers then met passengers at disembarkation points at LHR airport to collect completed questionnaires and consent passengers to follow-up interviews.

#### Semi-structured interviews

Passengers who consented to participate in a follow-up interview were contacted by email to confirm an interview time and language preference (English/Mandarin). Follow-up emails were sent out after 10 days if no response was received. After confirmation by participants, one-to-one telephone interviews of approximately 30 minutes were conducted between 2^nd^ and 23^rd^ April 2020.

During the interview, participants were asked about the COVID-19 information they received during their journey and their thoughts on the PHE information and advice provided. If interviewees had developed symptoms since arriving in the UK and were self-isolating/had self-isolated, they were also asked about their views and experiences of self-isolation using a separate topic guide.

All interviews were audio-recorded, and researchers created summaries of each interview. English interviews were transcribed *verbatim*; Mandarin interviews were transcribed directly into English.

## Data analysis

Categorical data were described as proportions and continuous data as median with interquartile range (IQR). All analyses were conducted in Stata v15.1 (2017, StataCorp LLC, College Station, TX).

Interview transcripts were coded initially using open coding. An initial coding framework was collaboratively developed by four researchers (TZ, SC, CS, WR) each coding one of the interviews they had conducted. Memos on emerging ideas and possible relationships between codes were kept alongside initial codes, and codes that represented similar concepts were assembled into conceptual categories and themes. Two (TZ, SC) of the research team then used the codes and categories in the framework to index each transcript in NVivo 12 Pro. Coding was performed iteratively within and between transcripts; common categories emerged across the transcripts, indicating data saturation.[7]

### Ethics statement

This study was a form of service evaluation, therefore no ethical approval was required. This was confirmed by PHE’s ethics committee - PHE Research Ethics and Governance Group.

## RESULTS

### Stage I – PPI interviews

Our PPI interviews included seven Chinese participants, five university students and two full-time employed staff, who had flown into the UK between 15^th^ January and 7^th^ February. All listed cough, difficulty breathing and fever as COVID-19 symptoms and stated that they would stay at home and call NHS 111 if they were symptomatic. In terms of official advice at that time, most participants believed the advice focusing on people travelling from Wuhan/Hubei was redundant following the compulsory lockdown in Wuhan. Conversely, the guidance on self-isolation and calling NHS 111 was seen as sensible and good advice; two participants (both employed) had voluntarily self-isolated when arriving in the UK as a precaution despite having no symptoms. Interviewees also expressed concerns about barriers to following this advice, such as important exams that students must attend, difficulties isolating from other people in a shared house, and potential language barriers to contacting NHS 111.

### Stage II – Airport cross-sectional survey and follow-up interviews

#### Survey results

##### Demographic characteristics

In total, 121 completed questionnaires from passengers on three flights operated by different airlines (Singapore Airlines, Air China and China Eastern Airlines) were collected. While 121 questionnaires were completed to a standard sufficient for inclusion, there was still some missing data due to people skipping some questions, putting prefer not to say, or illegible writing. Of those who answered (n=117), the age range was 20 to 81 years (median 53, IQR 36–64 years); 48/120 (40.0%) were male and 72/120 (60.0%) female. Just over half of respondents were British (n=64/118; 54.2%), 25.4% (n=30/118) were Chinese and 20.3% (n=24/118) ‘Other’. The majority of respondents could read English fluently (n=99/118; 84.0%), 14 were bilingual and 4 trilingual. There were 17 (14.4%) who could only read Mandarin and 1 (1.0%) who could only read Cantonese. None of the respondents had been to Wuhan city of Hubei province in mainland China in the 14 days prior to arriving at LHR.

##### Knowledge of symptoms and actions to take

The majority of respondents correctly identified a fever/high temperature (87.6%), difficulty breathing (87.6%) and cough (85.1%) as the symptoms associated with COVID-19 (Table 1). In line with the official case definition at the time (described in the PHE leaflets as cough, fever or shortness of breath), 101 (83%) of 121 respondents identified all three symptoms as symptoms of COVID-19. Other symptoms not included in PHE’s advice were also reported by passengers as COVID symptoms including commonly fatigue (57.9%), sore throat (52.9%) and headache, chills/shivering, aching limbs, runny nose and sneezing (>40%).

**Table 1.**
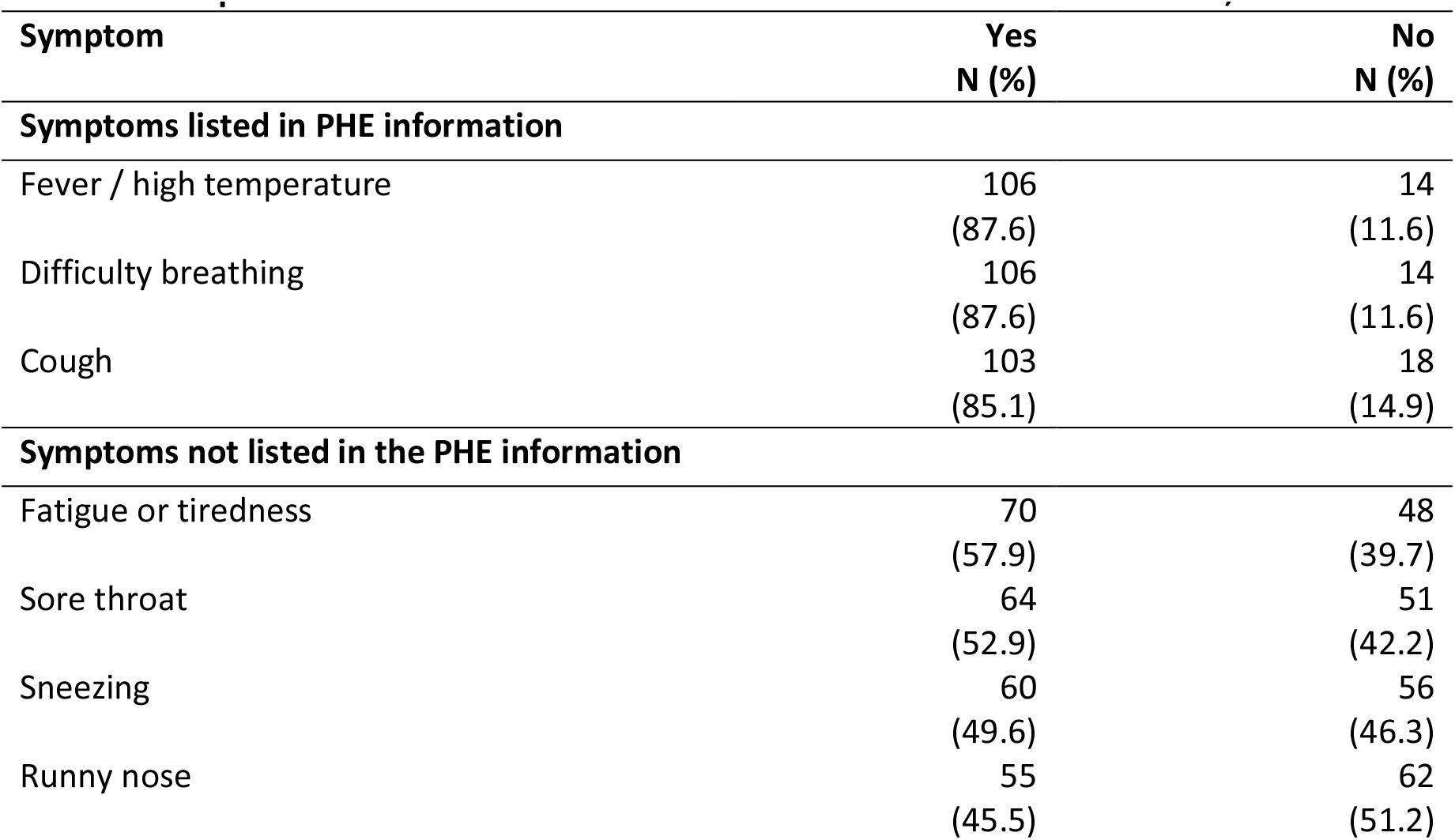
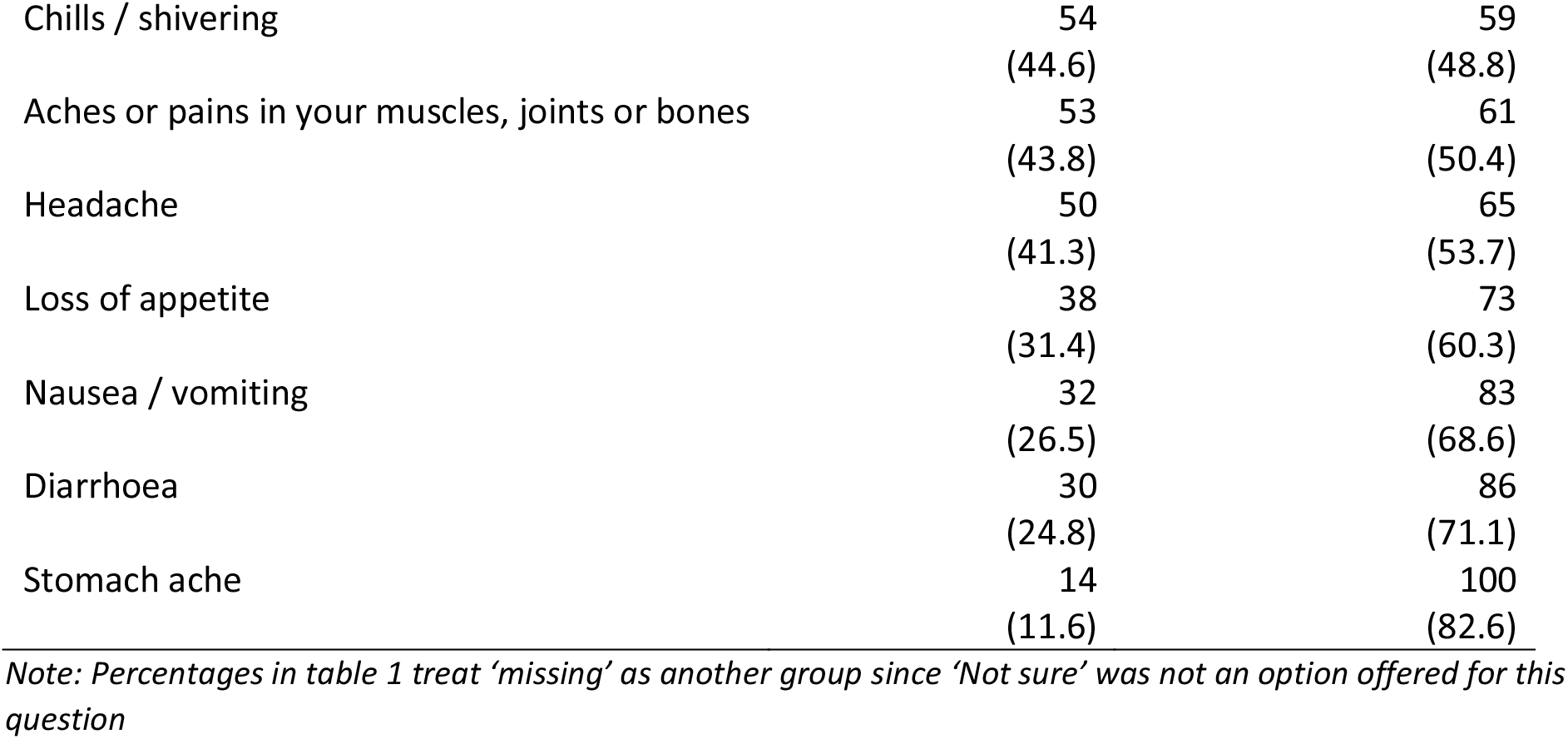
Recognition of COVID-19 symptoms in a sample of 121 passengers arriving at London Heathrow airport from COVID-19 affected countries between 4^th^ and 13^th^ March, 2020.

Most participants were correctly aware that people with COVID-19 might not show symptoms straight away (77.1%) and that asymptomatic status could last for 14 days (75.4%). 92.4% of participants also thought that people with COVID-19 can be contagious, even if they did not develop any symptoms. A minority of respondents (9.3%) mistakenly thought antibiotics could treat COVID-19 but a substantial proportion (27.1%) were uncertain about this.

Table 2 shows that most passengers were able to identify the recommended actions to take if they had been to Wuhan in the previous 14 days – to self-isolate (96.6%) and call NHS 111 for advice (84.6%). Respondents were less confident about actions to take for those who had been to other highlighted destinations (Box 1, versions 3 & 4); For people who had travelled to Singapore in the past 14 days, the majority correctly stated that they should not take any action if well, in accordance with PHE information; however, a substantial minority thought they should self-isolate (23.7%) and call NHS 111 for advice (18.8%), respectively, while the PHE leaflets advised these actions only for those with symptoms.

**Table 2.**
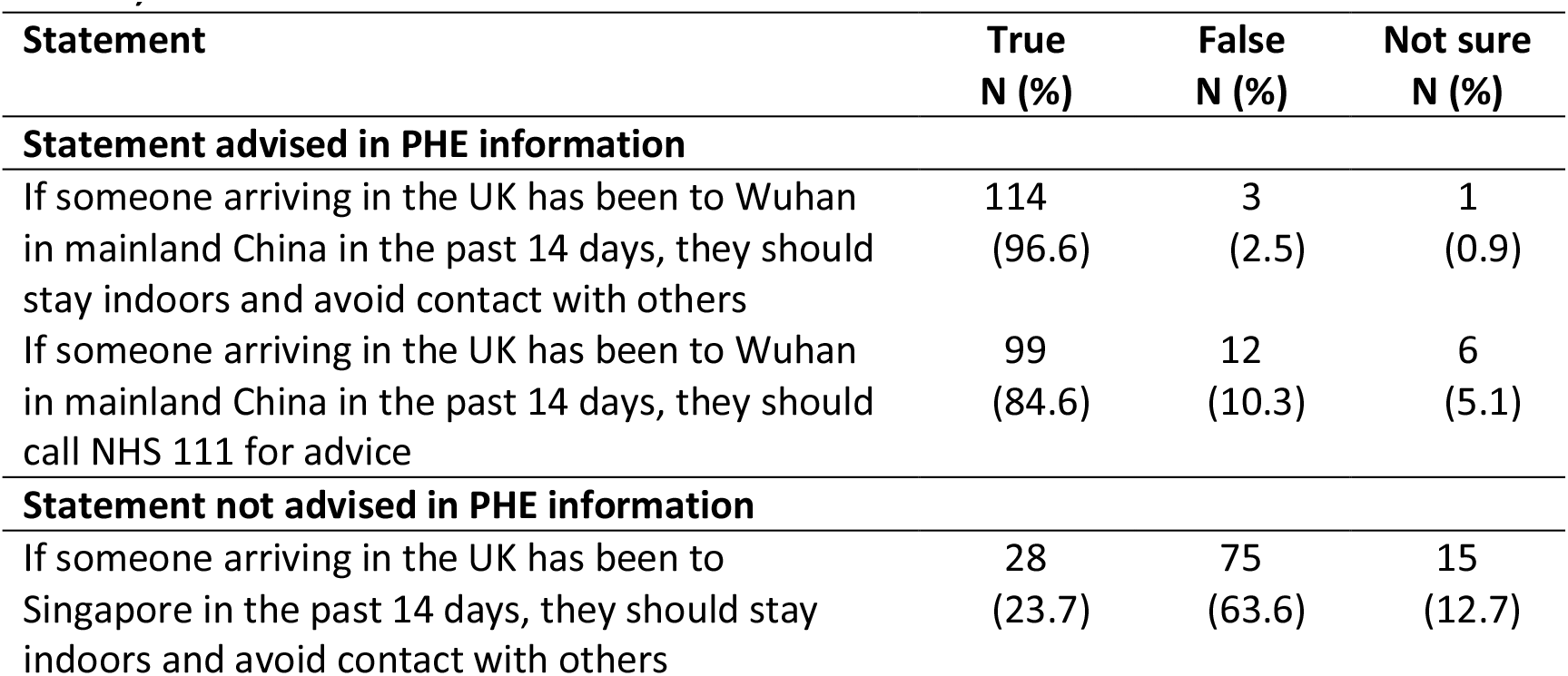
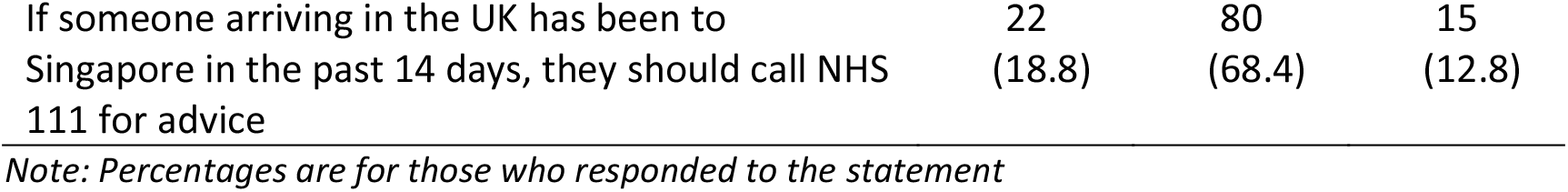
Knowledge of health-seeking behaviour in a sample of 121 passengers arriving at London Heathrow airport from COVID-19 affected countries between 4th and 13th March, 2020.

| Statement | True<br>N (%) | False<br>N (%) | Not sure<br>N (%) |
| --- | --- | --- | --- |
| **Statement advised in PHE information** |  |  |  |
| If someone arriving in the UK has been to Wuhan in mainland China in the past 14 days, they should stay indoors and avoid contact with others | 114<br>(96.6) | 3<br>(2.5) | 1<br>(0.9) |
| If someone arriving in the UK has been to Wuhan in mainland China in the past 14 days, they should call NHS 111 for advice | 99<br>(84.6) | 12<br>(10.3) | 6<br>(5.1) |
| **Statement not advised in PHE information** |  |  |  |
| If someone arriving in the UK has been to Singapore in the past 14 days, they should stay indoors and avoid contact with others | 28<br>(23.7) | 75<br>(63.6) | 15<br>(12.7) |
| If someone arriving in the UK has been to Singapore in the past 14 days, they should call NHS 111 for advice | 22<br>(18.8) | 80<br>(68.4) | 15<br>(12.8) |

*Note: Percentages are for those who responded to the statement*

##### Attitudes to official advice

In total 104/121 (86.0%) passengers stated that they had read the leaflet (94 read the English version, 30 read the Mandarin version and 20 read it in both languages). Only 6 (5.0%) stated that they had not read it in either language.

Overall, respondents thought the leaflet and poster at the airport were easy to understand (84.4% agree or strongly agree) and trusted the official advice (84.2% agree or strongly agree). In addition, the majority of respondents agreed that they had received enough information, including what to do if they developed COVID-19 symptoms (84.1%) and how and when to avoid contact with others (84.2%) (Table 3). Respondents may have reported their responses to the leaflet handed out on the flight, rather than to the leaflets and posters at the airport, but both had the same content.

**Table 3.**
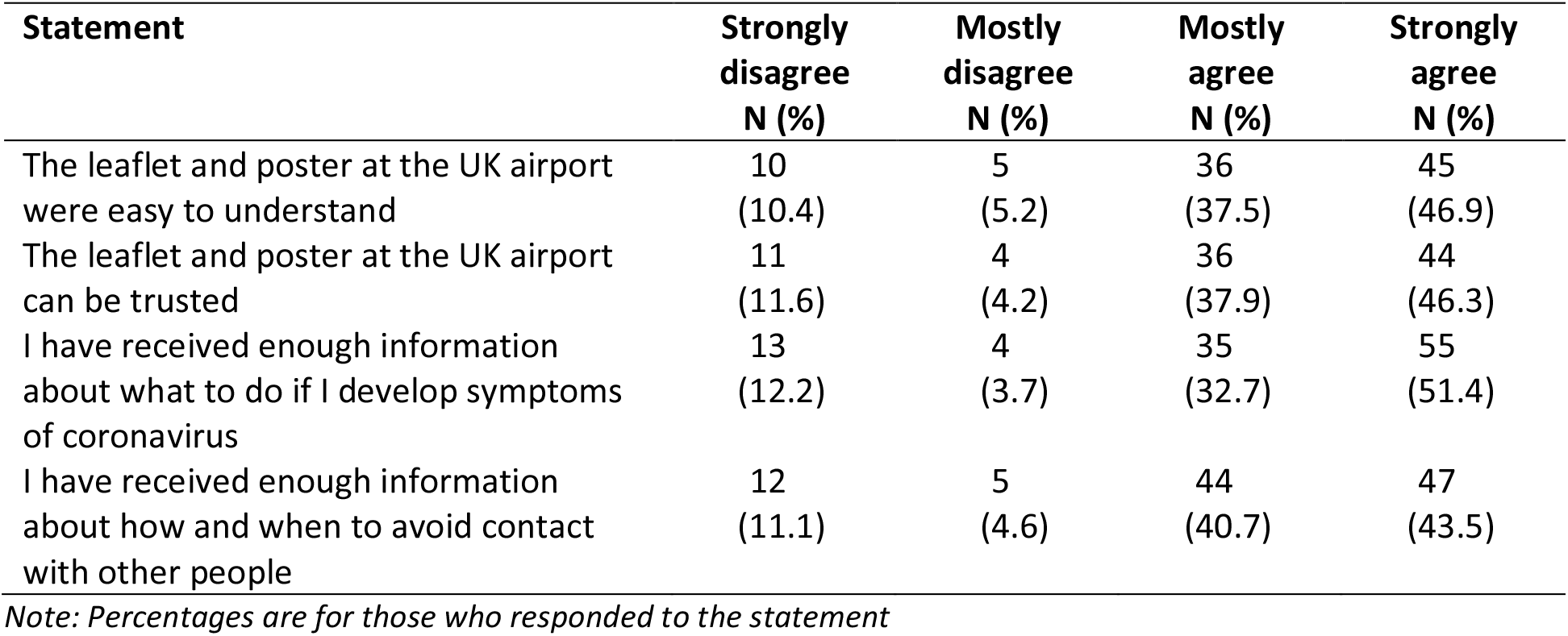
Attitudes to official Public Health England advice in a sample of 104 passengers arriving at London Heathrow airport from COVID-19 affected countries between 4th and 13th March, 2020.

##### Qualitative findings

15 interviews were conducted with passengers who had arrived on surveyed flights and consented to participate when completing the questionnaire. The age range was 21 to over 80 years, with five men and 10 women. Among the 15 participants, six were retired, five worked full-time, three were full-time students and one was unemployed. Most participants were permanent residents in the UK; three were limited-duration residents and two were temporary visitors. The majority (11 participants) were British, three were Chinese, and one was from New Zealand. All three Chinese participants could speak Mandarin and English and had seen PHE information in both languages. All White participants could speak only English.

The results represent passengers’ views and perspectives on the public health advice and their experience of self-isolation. These views clustered into five broad themes, (i) understandings related to COVID-19, including sub-themes of COVID-19 knowledge, personal or lived experience, exposure, domestic concerns, personal protective equipment; (ii) attitudes towards information materials and presence, self-isolation and lockdown, and sub-themes including attitudes to advice, information and presence, attitudes to self-isolation and lockdown, public adherence and perceptions of others, social pressure; (iii) practices and experience during the pandemic, and sub-themes of difficulties, feeling lucky, self-disciplinary, compulsory measures; (iv) Information and advice, and sub-themes of UK official advice, other source information, clear, reliability; (v) Support, and sub-themes including emotional support, healthcare support, information support, instrumental support. Only those themes relating directly to the reception of public health advice are reported below.

##### Knowledge of symptoms and actions to take if symptomatic

13 out of 15 participants recalled that they received the information leaflet from UK authorities/PHE during the flight or at the airport in Singapore or China. Most were impressed with the amount of information and measures being taken at departure airports and surprised that *‘there was almost nothing*’ [Participant 11] and ‘*nobody seemed to care*’ [Participant 8] on arrival at LHR. Only three passengers saw posters, which they said were not eye-catching, and two mentioned there was hand sanitiser at passport control area:

*‘I walked fast passing (those leaflets/posters), didn’t pay much attention*.*’* [Participant 15]

Cough, fever/high temperature and, as the disease progresses, breathing difficulties were the most frequently mentioned symptoms; ‘*you may be asymptomatic and so you have a cough or you might come down with a full-blown fever to the point where really you cannot breathe*’ [Participant 1].

Many passengers believed that other diverse symptoms such as headache, fatigue, loss of smell and taste were associated with COVID-19 but they were not included in the official case definition (see Table 1) at the time.

Most participants noted that they would start with self-isolation when symptoms were mild, and NHS 111 should be called if symptoms progress, indicating they would follow the official advice and base their actions on the disease severity:

‘*Well the first thing I would have had to have done would be to self-isolate. … And if the symptoms obviously got progressively worse I would then either contact my GP or phone 111. But it’s a fairly straightforward process that’s been set up to do this*’. [Participant 9]

##### Attitudes to official advice

The content of UK official advice itself was seen as reasonable and adequate; the majority of passengers commented that it was ‘*quite clear and sensible*’ [Participant 5] and felt the government was taking some action in response to the outbreak. Some participants believed more information or further clarification of the official advice should be given; for example, one passenger who stayed in the UK for nine days suggested adding a message on the official leaflet that, ‘*if you are experiencing any symptoms of coronavirus please contact your airline company and do not travel*’ [Participant 1].

Participants mentioned common concerns that people may disregard official advice in the UK, citing their lived experience in affected countries where televised public health information for COVID-19, including on social distancing and washing your hands, was ‘*reinforced every time there was a commercial break*’, whereas in the UK ‘*it’s random*’ [Participant 2]. They noted that the lack of visible pandemic control measures at LHR gave ‘*a false sense of security*’ [Participant 7] and therefore suggested reinforcing official advice and taking other actions such as installing temperature scanners, handing out materials and increasing the number of personnel at airports, as well as enacting more compulsory measures and regulations to limit close contact and quarantine arrivals.

‘*At Heathrow, we arrived and it was like nothing was wrong in the UK, so I think that causes a false sense of security, so maybe if there was more of a presence, like information, temperature check, personnel etc, people might take it more seriously*.’ [Participant 5]

‘*Well they could have had thermal imaging cameras, they could have had medical staff in protective clothing there to talk to people whose temperature came up as above the norm, they could have then asked people in those conditions, you know, if they met those conditions to isolate them, you know*.’ [Participant 8]

##### Acting on official advice

Most participants had acquired information and advice from both the UK and the countries of departure, regardless of their usual country of residence. Since COVID-19 had already spread across the countries from which they had travelled, participants believed they were ‘*educated enough about it*’ [Participant 7] and that they treated it more seriously than people in the UK; they were, as one participant put it, ‘*a bit ahead of the game*’ [Participant 3]. When arriving in the UK, as a precaution many participants voluntarily self-isolated or tried to distance themselves and skipped certain activities where people would be gathering, even though this was not the official advice at that time:

‘…*even though there wasn’t the, you know, that wasn’t really about the distancing over here, but we just thought we won’t see family and friends for some time just because we’d been or gone through Singapore*.’ [Participant 3]

‘*I didn’t dare go to the university to take the exam on Monday, because the teacher said if you didn’t feel well you could stay at home and didn’t have to go to the university to take the exam*.’ [Participant 13]

Participants expressed awareness of their risk of exposure while travelling that led some of them to self-isolate. They further noted that by doing so, they would not get blamed if any of their loved ones did get sick; one said they knew there was likely going to be a ‘stigma’ around them since they were coming from an affected country [Participant 10]: ‘…*but being on the plane with other people coming from who knows where with who knows what, you know, we were a bit more concerned which is why we isolated when we came home*.’ [Participant 11]

‘*We didn’t want to put any of our family members or friends at risk in case we were carrying the virus but didn’t know it*.’ [Participate 2]

*‘…we sort of knew pretty much that the chances of us giving him (family member) anything were miniscule, because we wouldn’t have put anybody at risk if we really thought that there was a chance but we just didn’t want it on us’*. [Participant 3]

They observed that tasks needing to be done outside the home could affect how people act on the advice; they therefore suggested *‘the British supermarkets and the Chinese supermarkets can offer home delivery*’ [Participant 13], which could contribute to minimise daily population flow and interaction. Despite experiencing some mental pressure, participants expressed they felt fortunate to have the physical and social resources to manage their self-isolation effectively, while being aware that this was not the case for everyone:

‘T*here is a temptation for people to get back home before you go to see a doctor, particularly if you’re living in a… going to a foreign country, you’re going there on holiday*.’ [Participant 1]

‘*I can’t think about it, I have to think about we’re very lucky, we’re luckier than most and if I want to go down and walk along the beach I sort of can. … I think if somebody is locked up in a one-bedroom flat in London it will be horrible, it must be horrible for them…*’ [Participant 10]

The reasonable and clear official information was seen to shape the public’s authentic understanding of the COVID-19 crisis and could therefore promote public acceptance of official advice. Participants further emphasised the crucial role of community support; ‘*I think providing they have sufficient support in their communities there is no reason at all why anybody should not self-isolate*’ [Participant 9].

‘*I can’t think why you would not follow the official advice but I think the mere…at the time the number of people who had died from Coronavirus it was rising but I think… and the numbers were unclear, but they were talking about one to two percent of the people who got infected may die…*’ [Participant 1]

## DISCUSSION

Our findings show that passengers arriving from China and Singapore during the early stages of the COVID-19 pandemic in the UK found the content of official public health information provided in PHE leaflets to be clear and easy to understand. Most could correctly identify the recommendations concerning actions to be taken in the event of becoming symptomatic/arriving from certain destinations and considered the advice provided to be acceptable and trustworthy. However, there was some uncertainty as to whether those arriving from one of those countries or territories listed in PHE information other than Hubei or Wuhan should proactively self-isolate or call the NHS helpline. The majority of those surveyed (83%) and interviewed were also able to correctly identify all three symptoms described in the leaflets and poster (cough, fever, shortness of breath), but over half of those surveyed and many of those interviewed also identified fatigue and sore throat as symptoms, with substantial proportions identifying other symptoms that were not included in the official case definition during the evaluation period. This definition changed over time, alongside evolving scientific knowledge of the virus, and some symptoms identified by our respondents had been recognised as the common manifestations of COVID-19, including anosmia that has more recently been introduced into the official PHE case definition. Since these passengers were arriving from countries where the pandemic, as well as COVID-19 policy responses to it, were further developed than in England when the study was conducted,[8, 9] their responses may well reflect knowledge acquired elsewhere as much as the official PHE advice.

Support for this is shown by the fact that while most survey respondents indicated they had received sufficient information both about what to do if symptoms developed and about how and when to avoid contact with other people, the PHE leaflets and posters provided no information on avoidance of contact, beyond the requirement to stay indoors if symptomatic or when arriving from specified source locations. Our interview data support the survey findings that respondents believed the official information was adequate; however, their accounts show that respondents’ knowledge was substantially informed by familiarity with public health interventions being taken elsewhere to contain transmission. On the other hand, for these passengers, the perceived lack of visible measures in place on arrival into the UK indicated a worrying lack of official concern about COVID-19. Their comments were verified by our research team’s observations that the design and positioning of PHE information at the arrival airport made it largely unnoticeable to arriving passengers (see Fig 1), as well as other studies that have pointed out an initial rejection of ‘eye catching measures’ in the UK at the beginning of the outbreak.[10] Such expressions of concern among passengers suggest that while the intended purpose of the leaflets was to provide information and guidance that would encourage people to act in accordance with recommended behaviours, recipients regard the provision of information and other visible public health measures as an index of the adequacy of governmental response to the pandemic. Therefore, the advice and information served two purposes during the pandemic – the original role of providing public health messaging, and also reflecting the performance of government and authorities. Under the circumstances that passengers have been well informed by knowledge from elsewhere, they seem to be more concerned about the other role, the performance, of advice and information. Respondents’ good understanding of the content of the PHE leaflet, which they received on the plane, contrasted starkly with their reports of low visibility of, and minimal interaction with, similar materials on arrival at the airport. This suggests that providing public health information in-flight, by announcements and distribution of leaflets – when passengers have the time to read it with few distractions – may be the more effective strategy.

**Figure 1.**
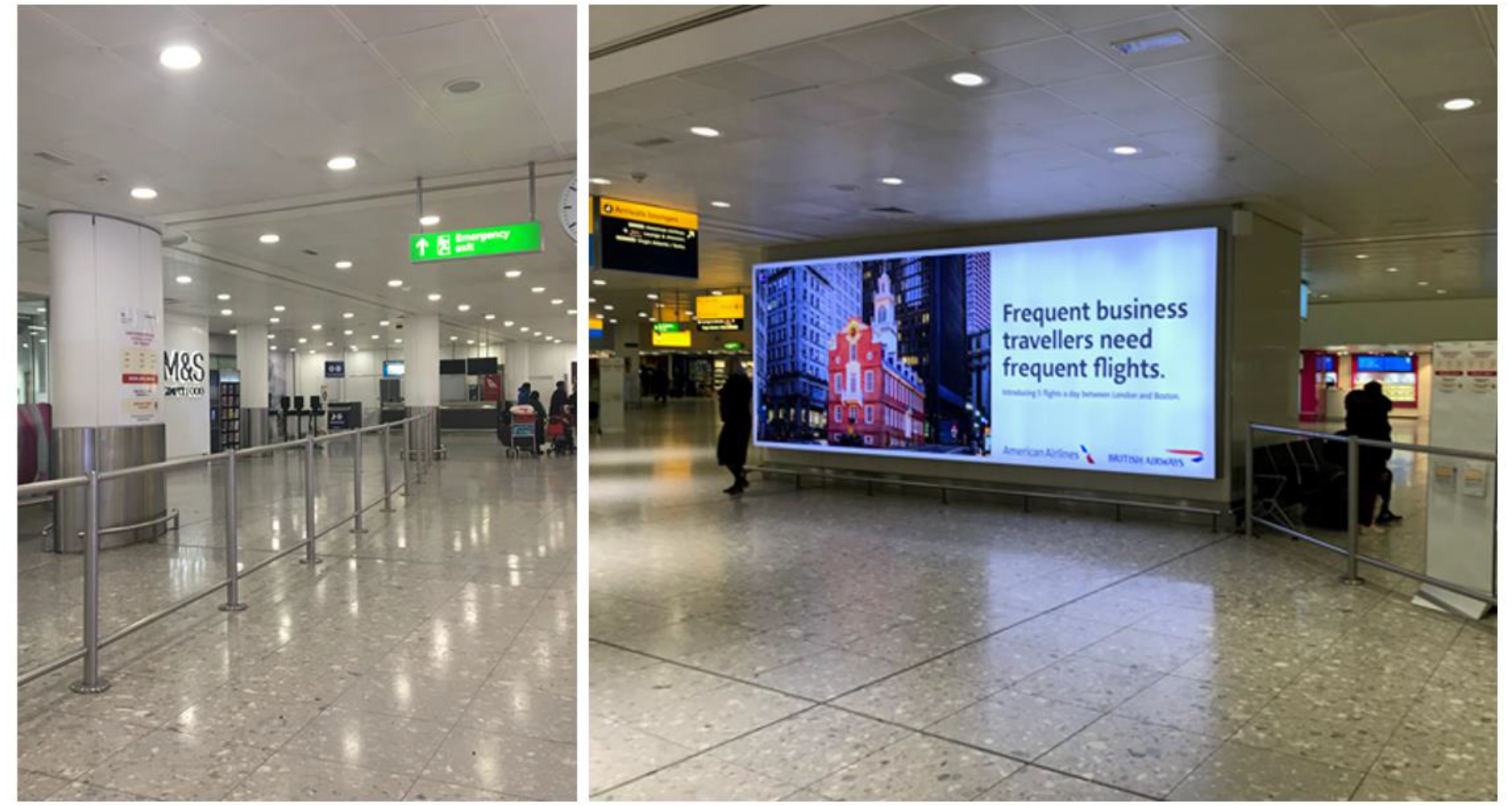
Public Health England poster, providing health and safety advice on COVID-19, at Terminal 3 Arrivals, London Heathrow Airport in west London, 4 March 2020. *Source: photos taken by researchers* *Note: left, COVID-19 poster on pillar; right, COVID-19 leaflet stand (right hand side)*

Our interviewees suggested a range of additional public health interventions that were not in force when they arrived, such as restricted contact tracing, widespread testing and self-isolation/quarantine for all arrivals, many of which were used for the purpose of containing viral transmission in countries like Singapore and China and were eventually implemented in the UK.[11] This again indicates that passengers were using prior experience of pandemic control measures elsewhere to judge the extent to which UK authorities were taking the pandemic seriously, based on their experience of arrival at a UK airport. The survey and interviewees’ accounts pointed to their own self-discipline not only in acting on official UK advice regarding self-isolation for those arriving from certain destinations or subsequently developing symptoms, but also in some cases going beyond it by doing spontaneously as an additional protective measure. This predominantly related to their perceptions of exposure risk in affected countries from which they had travelled or from the journey itself, as well as to concerns about potential stigmatisation should family or colleagues subsequently become infected. Similar findings have been reported by previous studies[12, 13] indicating that travellers arriving from Ebola-affected countries restricted movement to avoid stigmatisation by the community. The public health interventions mentioned by passengers and their behaviours suggest screening people at entry, as done in ‘enhanced screening’ for Ebola, may help to reassure the travelling public that containment measures are in place, even if border screening is in reality ineffective. However, screening measures generate other difficulties such as how to accurately identify targeted passengers and how to avoid social stigmatisation.[5, 14]

This study has a number of limitations. In our qualitative work, interviewees were recruited based on survey participants’ willingness to consent to follow-up interviews rather than from a purposive sample. It is possible that such participants may hold stronger views than those who refuse, or may come from particular groups such as retired people. The interviewees’ accounts may therefore not accurately represent the views of all passengers who completed the survey. However, the demographic characteristics of interviewees indicated a relatively comprehensive coverage of both British residents and other nationalities coming to the UK for a variety of purposes, so the sample composition seemed reasonable. Given the time that elapsed between survey completion and interview as well as influences from the rapid changes in both pandemic and UK policies, interviewees’ impressions on the presence at the airport and their perceptions and views might have changed in the interim; all interviews were completed within 7 weeks from passengers’ arrive date to minimise these effects. Finally, both study size and early opportunities to use our findings to inform the content and delivery of official public health guidance were limited by difficulties in gaining airside access at airports and obtaining cooperation from airlines, so that by the time we implemented data collection, the number of flights and passengers arriving from affected countries had already diminished drastically.

## CONCLUSION

Our findings confirm the clarity and acceptability of public health guidance on COVID-19 provided to passengers arriving into UK ports in the early stages of the pandemic. However, they also demonstrate a widespread perception that the provision of information alone on arrival was an insufficient official response to this global public health emergency at that stage in the pandemic. This finding is cause for concern since it may reduce trust in official sources, an established driver of nonadherence to public health interventions.[15] It also indicates that the public health information provision at borders should be appraised not only with respect to its functional effectiveness in imparting guidance and encouraging behaviours to control transmission, but also with regard to its perceived effectiveness in furnishing public assurance of official action to contain the disease threat. We also found that travellers arriving from countries where the epidemic was already established frequently had knowledge of COVID-19 and of various public health measures to contain transmission that were not derived from official UK advice or present in the UK at that time. In a rapidly evolving international health crisis, and particularly one in which understanding of the disease threat is partial and changing, evaluating public understanding by reference to locally defined parameters can be unreliable, not least because public knowledge among those with experience from elsewhere may be more advanced than local understanding. This indicates the value of appraising public perceptions not only to measure understanding and adherence, but also to gain potential insights into future appropriate measures and their likely acceptability. Our study also demonstrates the complexity of health policy decision-making in public health emergencies of international importance and provides fresh insights into the need to take greater account of the diverse information sources on which international travellers may draw. Finally, it highlights the importance of establishing efficient mechanisms for rapid appraisal and feedback to public health and regulatory authorities of evidence that could contribute to containment and control of epidemic disease threats.

## Data Availability

The data generated or analysed during this study are included in this article; data are available from authors from Public Health England or University of Bristol upon reasonable request

## ACKNOWLEDGEMENT

We wish to thank all participants who contributed to this study and all staff in London Heathrow Airport for their kind support. All authors contributed to, and read and approved, the final version of the manuscript. Charlotte Robin is affiliated to the National Institute for Health Research Health Protection Research Unit (NIHR HPRU) in Emerging and Zoonotic Infections at University of Liverpool in partnership with Public Health England (PHE), in collaboration with Liverpool School of Tropical Medicine and The University of Oxford, the NIHR HPRU in Gastrointestinal Infections at University of Liverpool in partnership with PHE, in collaboration with University of Warwick and the NIHR HPRU in Behavioural Science and Evaluation at University of Bristol, in partnership with PHE. Charlotte Robin is based at PHE. The views expressed are those of the author(s) and not necessarily those of the NHS, the NIHR, the Department of Health or Public Health England. Lucy Yardley is an NIHR Senior Investigator and her research programme is partly supported by NIHR Applied Research Collaboration (ARC)-West, NIHR Health Protection Research Unit (HPRU) for Behavioural Science and Evaluation, and the NIHR Southampton Biomedical Research Centre (BRC). For any enquiries about the data please contact Public Health England or University of Bristol.

